# Individualized Per-Site Meta-Federated Feature Learning (iPS-MFFL) for Privacy-Preserving Brain Tumor MRI Classification under non-IID Heterogeneity

**DOI:** 10.64898/2026.04.15.26351000

**Authors:** Yuto Hakata, Miko Oikawa, Shin Fujisawa

## Abstract

**Background:** Federated learning (FL) enables collaborative model training across institutions without sharing patient-level data. However, standard FL algorithms such as FedAvg degrade under non-independently and non-identically distributed (non-IID) data, a prevalent condition when patient demographics, scanner hardware, and disease prevalence differ across hospital sites.

**Objective:** We propose **iPS-MFFL (Individualized Per-Site Meta-Federated Feature Learning)**, a federated framework with a hierarchical local-model architecture that addresses non-IID heterogeneity through (1) a shared feature extractor, (2) **multiple weak-learner classification heads that can be trained with heterogeneous training objectives** to promote complementary decision boundaries, (3) **independent per-learner server aggregation** so that each weak learner’s parameters are averaged only with its counterparts at other clients, and (4) a lightweight meta-model — itself federated — that adaptively stacks the weak-learner outputs. The specific choices of backbone, weak-learner training objectives, and meta-model are implementation details; in this work we use an ImageNet-pretrained ResNet18 and three heterogeneous losses as a concrete instantiation.

**Methods:** We evaluate on the Brain Tumor MRI Classification dataset (7,200 images; 4 classes: glioma, meningioma, pituitary tumor, no tumor) partitioned across K = 5 simulated hospital sites using Dirichlet non-IID sampling (α = 0.3). Four baselines are compared: Local-only training, FedAvg, FedProx, and Freeze-FT. All experiments are repeated over three random seeds (13, 42, 2025) and evaluated using paired t-tests, Cohen’s d effect sizes, and post-hoc power analysis.

**Results:** iPS-MFFL achieved the highest mean final-round test accuracy point estimate of **85.42 ± 8.70%** (mean ± SD across three seeds), compared to FedAvg (78.48 ± 12.66%), FedProx (78.33 ± 14.64%), Freeze-FT (73.98 ± 21.09%), and Local (58.10 ± 11.77%). iPS-MFFL showed the smallest cross-seed SD, suggesting greater robustness to partition heterogeneity. However, one-way ANOVA did not reach statistical significance (F = 1.52, p = 0.270), reflecting the limited number of experimental seeds. Cohen’s d effect sizes relative to iPS-MFFL ranged from 0.59 (vs. FedProx) to 2.64 (vs. Local); post-hoc pairwise comparisons are reported as exploratory given the non-significant omnibus test. Post-hoc power analysis indicated that statistical power for FL baseline comparisons was only 0.10–0.12 for the observed effect sizes (d ≈ 0.6) at n = 3 seeds.

**Conclusions:** iPS-MFFL provides a practical approach to heterogeneous federated brain tumor classification by combining transfer learning, contrastive weak-learner diversity, and meta-learning. The framework demonstrated the highest mean accuracy and lowest variance across diverse data partitions. Validation with larger seed pools (≥ 10 seeds for 80% power), ablation studies, and external multi-center cohorts is needed to establish generality.

## 1. Introduction

Adult diffuse glioma is a class of primary brain tumors that, under the 2021 WHO Classification of Tumors of the Central Nervous System [1], is now reorganized on the basis of molecular markers such as IDH mutation status and 1p/19q co-deletion.

MRI-based analysis plays a central role in the diagnosis and treatment planning of these tumors [2, 3]: accurate characterization of tumor imaging phenotypes is essential for determining the extent of surgical resection, defining target volumes in radiotherapy, and performing longitudinal assessment of treatment response [3, 4]. In particular, for glioblastoma (IDH-wildtype; WHO CNS grade 4) the median survival under standard temozolomide-based chemoradiotherapy remains approximately 14.6 months [5], and precise characterization directly impacts prognosis. The experimental validation in this work uses a 4-class brain tumor classification task (glioma, meningioma, pituitary tumor, and no tumor) and the findings should be interpreted within that scope.

Recent advances in deep learning have enabled automated analysis of brain tumor MRI, with convolutional neural networks (CNNs) achieving high accuracy on centralized datasets [6]. However, in clinical practice, brain tumor imaging data are distributed across multiple hospitals, each subject to privacy regulations such as HIPAA and GDPR. Centralizing patient data for model training is therefore often infeasible.

Federated learning (FL) [7] addresses this challenge by training a shared model collaboratively while keeping patient data on-site. In the canonical FedAvg algorithm [7], each site trains locally and sends only model parameters to a central server for weighted averaging. Despite its appeal, FedAvg degrades under **non-IID data** — when local distributions differ across clients [8]. In medical imaging, non-IID conditions arise naturally from differences in patient demographics, disease prevalence, imaging protocols, and scanner hardware.

Various approaches have been proposed to address non-IID heterogeneity, including proximal regularization [9], contrastive representation alignment [10], and decoupled feature-classifier training strategies [11]. However, existing methods typically rely on a single classification head, limiting their capacity to capture diverse aspects of the data distribution.

We argue that (1) leveraging a **pretrained deep backbone** for richer feature extraction, (2) maintaining **multiple weak learners** with heterogeneous training objectives to promote complementary decision boundaries, and (3) combining them with a **meta-model** for adaptive stacking can better exploit the complementary strengths of global and local knowledge.

**Why this combination?** The theoretical motivation for iPS-MFFL rests on three principles. First, ensemble diversity theory [14] establishes that an ensemble’s error decreases as the correlation between individual learners decreases; training weak learners with heterogeneous loss functions (cross-entropy, feature-contrastive, output-contrastive) directly reduces this correlation by inducing distinct error patterns. Second, freezing the backbone during fine-tuning creates a natural separation between representation learning (global) and decision boundary learning (local); replicating the fine-tuning step across multiple heads multiplies the diversity benefit without additional communication cost for the backbone. Third, meta-learning provides a principled mechanism for learning the optimal combination weights that adapt to each client’s data distribution, avoiding the need for hand-tuned aggregation rules.

Contributions:

1. **A federated learning framework for multiple weak learners**, in which each client holds a shared feature extractor together with N ≥ 2 weak-learner classification heads.
2. **Independent per-learner server aggregation** — each weak learner’s parameters are collected and averaged at its own dedicated aggregation server, so that the distinct characteristics of different weak learners are preserved rather than averaged out in a single pooled aggregation.
3. **Federated meta-model** — a lightweight meta-model that stacks the weak-learner outputs is itself aggregated at a separate server, allowing the combination strategy to reflect each client’s data distribution while still benefiting from cross-client knowledge sharing.
4. **A concrete instantiation** on a brain tumor MRI classification task, using an ImageNet-pretrained ResNet18 backbone, three weak learners trained with heterogeneous loss functions, and a small MLP meta-model, evaluated with multi-seed statistical testing and post-hoc power analysis.

## 2. Methods

### 2.1 Study design and reporting

This is a retrospective methodological investigation using a publicly available, anonymized brain tumor MRI classification dataset. It does not involve recruitment of new patients, any intervention, or handling of identifiable personal data.

### 2.2 Dataset

We use the **Brain Tumor MRI Classification** dataset publicly redistributed on Kaggle by Nickparvar [12], which is an augmented/curated collection derived from earlier publicly available brain tumor MRI sources including the dataset originally released by Cheng et al. [16]. The version used in this study contains 7,200 brain MRI images across four classes:

Images are grayscale JPEG at 512 × 512 pixels, resized to 128 × 128 for training. Standard data augmentation (random horizontal flip) and normalization (mean = 0.5, std = 0.5 per channel) are applied.

We note that this dataset, while widely used in FL research, is a community-curated Kaggle resource rather than a controlled clinical trial dataset. Its original provenance and curation methodology are not fully documented, which limits the clinical generalizability of findings derived from it (see Limitations, Section 4.7).

### 2.3 non-IID data simulation

To simulate cross-institutional statistical heterogeneity, we employ Dirichlet-based partitioning [13]. Given K = 5 clients and concentration parameter α = 0.3, the class proportions for each client are sampled from Dir(α), producing moderately to substantially skewed distributions (lower α yields more extreme heterogeneity; α = 0.1 is commonly labeled “highly non-IID” while α = 1.0 is near-uniform in standard FL benchmarks [13]). Each random seed generates a different partition with varying degrees of heterogeneity.

To quantify partition difficulty, we compute the **mean per-client Shannon entropy** (base 2) of the class distribution. Higher entropy indicates more balanced local distributions (maximum = 2.0 for 4 classes); lower entropy indicates more extreme heterogeneity.

The resulting partitions are visualized as heatmaps in **Figure 1**.

**Fig. 1.**
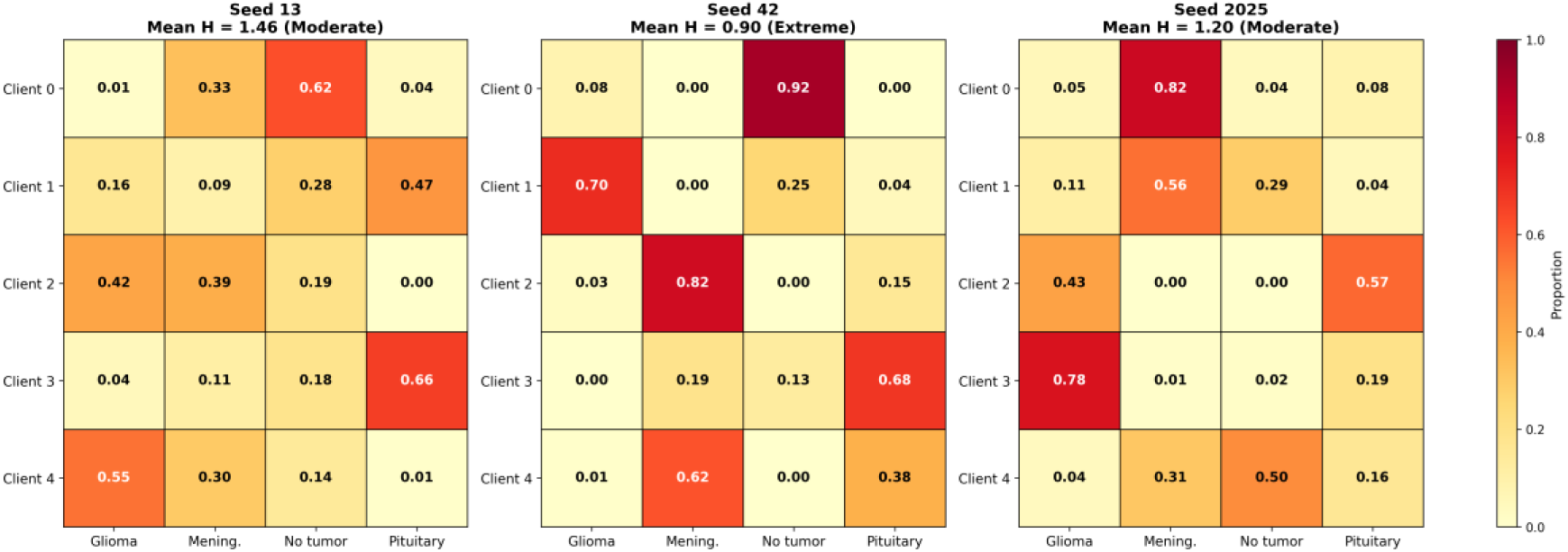
Non-IID data partitions across three random seeds (Dirichlet α = 0.3, K = 5 clients). Cell values show sample counts; color intensity represents class proportion. Mean per-client Shannon entropy (H̅) quantifies partition difficulty: seed 42 (H̅ = 0.91) produces the most extreme heterogeneity, with three zero-class instances.

Seed 42 produces the most challenging partition, with a mean client entropy nearly half the maximum and three instances where a client receives zero samples of a class. This creates a realistic worst case in multi-institutional medical imaging, where some clinics may handle a narrow range of conditions.

### 2.4 Model architecture (general framework)

Each client device holds a model composed of three abstract modules, illustrated in **Figurer 2**.

**Fig. 2.**
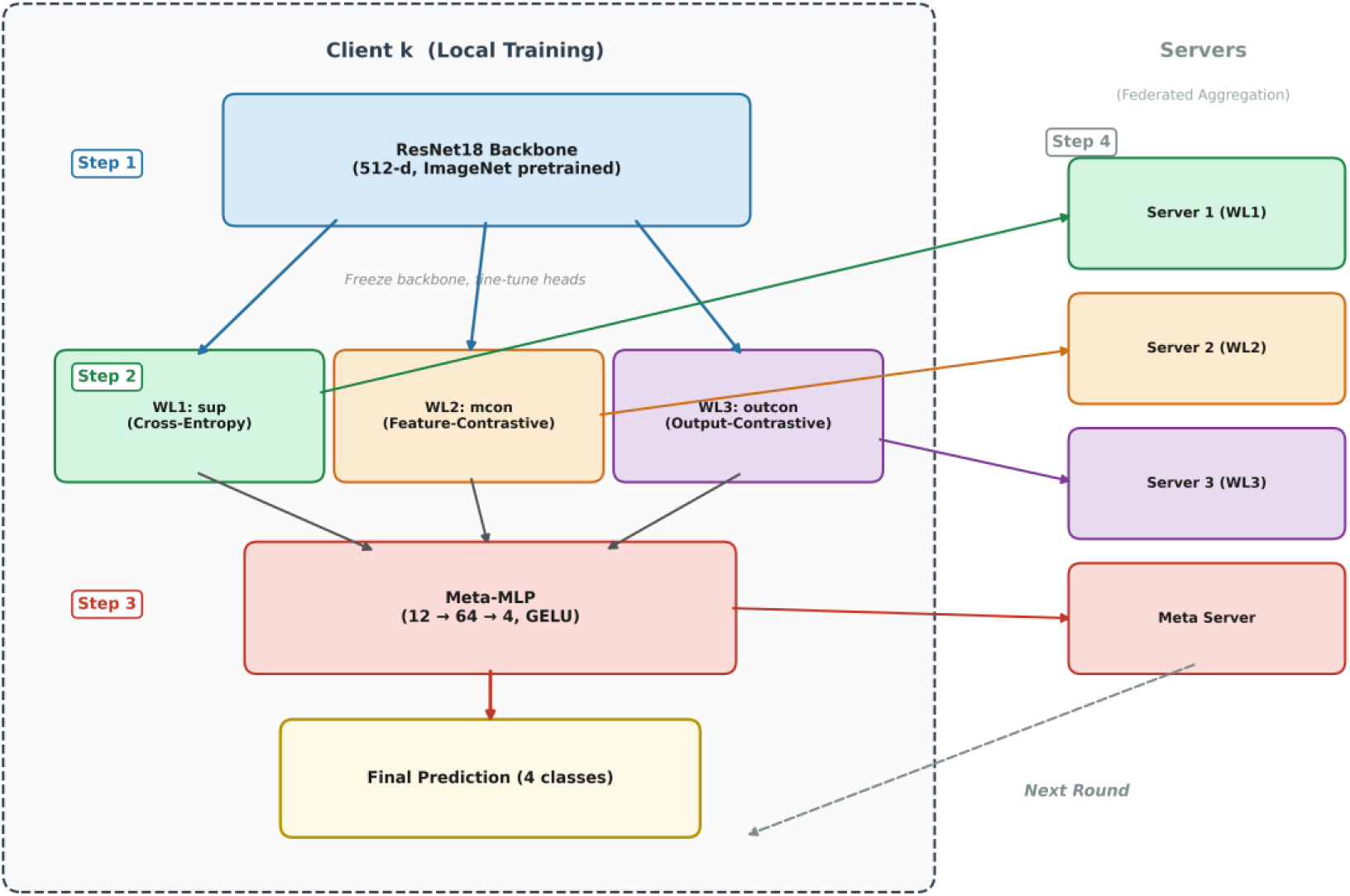
Overview of the iPS-MFFL architecture. Within each client, Step 1 trains the shared feature extractor jointly with the weak-learner heads. In Step 2, the feature extractor is frozen and the N weak-learner heads are fine-tuned independently. In Step 3, a meta-model combines the weak learners’ outputs. Step 4 aggregates each weak learner at its own dedicated server and the meta-model at a separate server.

**Fig. 2**. Overview of the iPS-MFFL framework with a hierarchical local-model architecture. The feature extractor, the N weak-learner heads, and the meta-model are each aggregated at independent servers. The specific choices of module types and training objectives used in this study are described in §2.7.

**Feature extractor (shared backbone).** A neural network *f*θ(·) that maps the input data to a fixed-dimensional feature vector. In this work we instantiate *f*θ as an **ImageNet-pretrained ResNet18** [6] producing a 512-dimensional feature vector (see §2.7 for implementation details).

**Weak-learner heads.** N ≥ 2 **independent classification heads** *g*1(·),…, *g*N(·) that share the feature extractor but **may be trained with different training objectives** to promote diverse (weakly correlated) decision boundaries. The framework is agnostic to the specific choice of training objectives; Ensemble theory [14] predicts that combining weakly correlated classifiers yields lower ensemble error than any individual classifier, motivating the use of heterogeneous objectives that induce decorrelated errors. The concrete objectives used in this study are described in §2.7.

**Meta-model.** A small trainable module *m*(·) that takes the outputs of the N weak learners as input and produces the final prediction. In this work we use a two-layer MLP; other choices (e.g., attention-based stacking, gating) are possible within the same framework.

**Servers.** The key structural feature of iPS-MFFL is that the model is federated via a hierarchy of **independent aggregation servers**: one server for the feature extractor, N servers for the N weak-learner heads, and one server for the meta-model. Each server collects only its corresponding parameters from the clients and aggregates them independently of the other servers. This preserves the distinct characteristics of different weak learners, which would otherwise be averaged out under a single pooled aggregation.

### 2.5 Training protocol

Training uses SGD with learning rate 0.01, momentum 0.9, and weight decay 1 × 10⁻⁴. Batch size is 256. Each federated round consists of four steps at every participating client:

**Step 1 — FedAvg base training.** The shared ResNet18 backbone and a base classifier are trained jointly using cross-entropy loss for 2 local epochs, establishing a shared feature representation starting from ImageNet-pretrained weights.

**Step 2 — Per-head fine-tuning.** The backbone is **frozen**. Each of the three weak-learner heads is independently fine-tuned with its specific loss function for 2 fine-tuning epochs. This step allows each head to specialize differently while preserving globally learned features.

**Step 3 — Meta-model training.** All backbone and weak-learner parameters are frozen. The meta-model is trained for 2 epochs on the stacked logits, learning the locally optimal combination for the client’s data distribution.

**Step 4 — Independent server aggregation.** The server performs weighted averaging (weights proportional to client data size) for each component independently: (1) each weak learner’s full model (backbone + head) at its own dedicated server, and (2) the meta-model separately.

### 2.6 Baselines

We compare iPS-MFFL against four baselines, all using the same ResNet18 backbone, optimizer settings, and training epochs:

1. **Local:** Each client trains independently without any communication. Lower bound on federated benefit.
2. **FedAvg** [7]: Standard federated averaging at a single server.
3. **FedProx** [9]: FedAvg with proximal regularization (μ = 0.01).
4. **Freeze-FT** [11]: After Step 1, the backbone is frozen and a single classification head is fine-tuned using cross-entropy loss. All parameters are aggregated at a single server.

**Note on Local baseline.** Local training runs for only 1 round (2 local epochs) because, without federated communication, additional rounds are equivalent to continued local training. The reported Local accuracy is evaluated by averaging the predictions of all K = 5 independently trained local models on the shared global test set (1,600 images), reflecting the performance achievable without any inter-site collaboration.

### 2.7 Concrete instantiation in this study

For the experiments reported in this paper, the abstract modules of §2.4 are instantiated as follows. We emphasize that these are implementation choices for the present study and do not delimit the scope of the framework.

- **Feature extractor:** ResNet18 [6] pretrained on ImageNet, with the final fully connected layer replaced by an identity mapping, producing a 512-dimensional feature vector. Input images are resized to 128 × 128 and converted to 3 channels.
- **Number of weak learners:** N = 3. Each weak learner is a linear layer (512 → 4).
- **Weak-learner training objectives:** Three heterogeneous objectives are assigned one-to-one to the three weak learners.
- `o` *WL1:* standard cross-entropy loss (supervised baseline).
- `o` *WL2:* cross-entropy plus a **feature-space contrastive term** that pulls the local feature representation toward that of the global model and pushes it away from that of the previous-round local model.
- `o` *WL3:* cross-entropy plus an **output-space contrastive term** that applies the same contrastive principle at the logit level.

These three objectives were chosen to induce decorrelated errors: WL1 supplies the pure discriminative signal, WL2 regularizes the feature space, and WL3 regularizes the output space. Other choices of N and of the per-learner training objectives are compatible with the framework.

- **Meta-model:** a two-layer MLP (3 × 4 = 12 → 64 → 4) with GELU activation that takes the concatenated logits of the three weak learners as input and produces the final four-class prediction.

### 2.8 Experimental setup

All experiments run for **10 federated rounds** with full client participation. Each experiment is repeated with three random seeds (13, 42, 2025) that control both the Dirichlet partition and model initialization. The held-out test set (1,600 images) is shared across all methods.

### 2.9 Evaluation and statistical analysis

Classification performance is evaluated using **overall accuracy** on the held-out test set. We report:

- **Mean ± standard deviation** of final-round and best-round test accuracy across three seeds.
- **Paired t-tests** (two-tailed, df = 2) between iPS-MFFL and each baseline.
- **Cohen’s d** effect sizes [15]. Benchmarks: d < 0.2 (negligible), 0.2–0.5 (small), 0.5–0.8 (medium), d > 0.8 (large).
- **Post-hoc power analysis** using the non-central t-distribution to determine the probability of detecting effects of the observed magnitude at n = 3.
- **One-way ANOVA** across all methods for omnibus group comparison.

We emphasize that n = 3 seeds provides severely limited statistical power. A paired t-test with df = 2 has a critical value of t = 4.303 (two-tailed, α = 0.05), requiring unrealistically large effect sizes to reach significance. We therefore report effect sizes as the primary measure and provide the required sample sizes for future studies targeting 80% power.

## 3. Results

### 3.1 Main results

These results are visualized in **Figure 3** (bar chart with individual seed points) and **Figure 5** (robustness analysis).

**Fig. 3.**
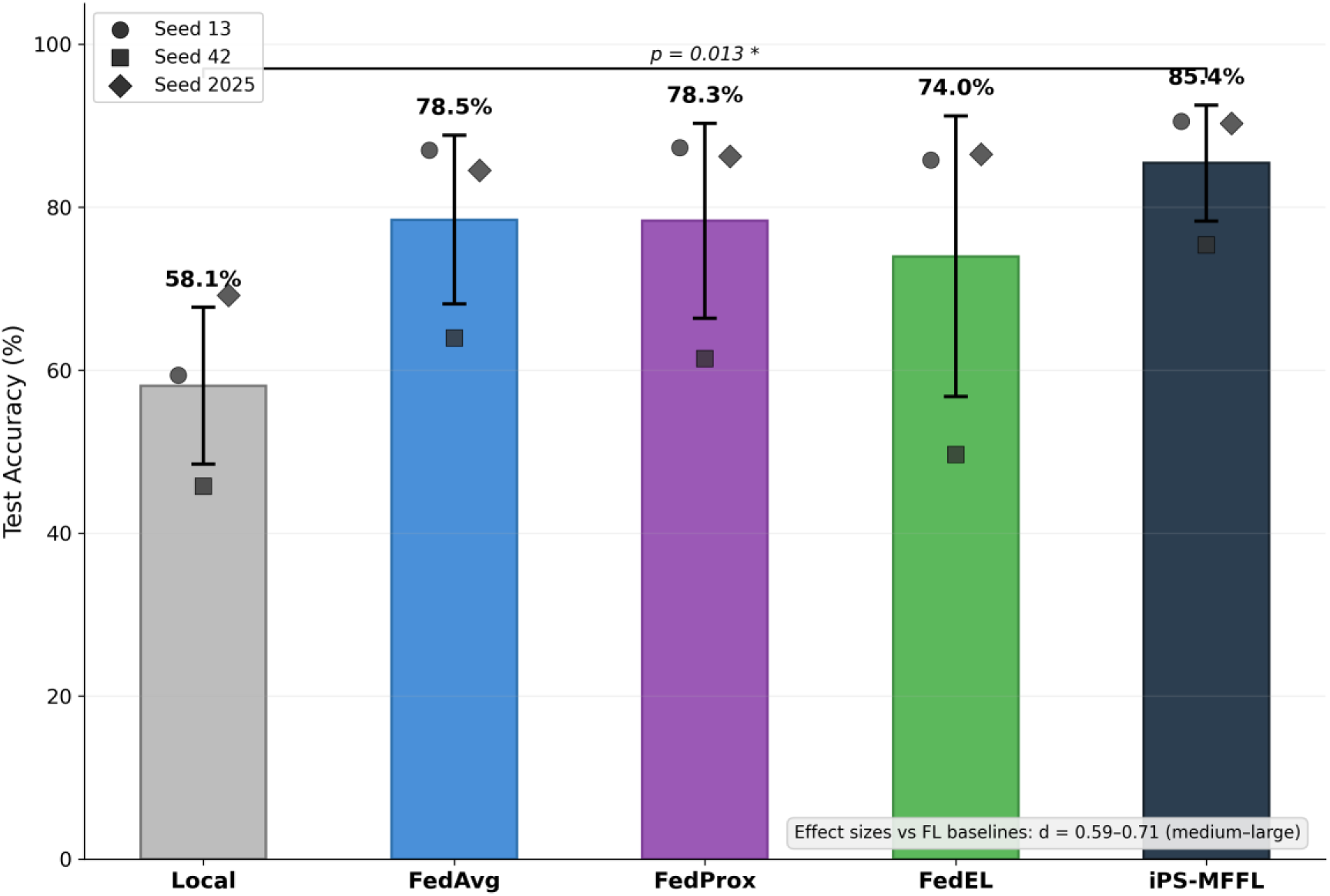
Final-round test accuracy (mean ± SD) across five methods and three seeds. Individual seed results are shown as scatter points (circle: seed 13, square: seed 42, diamond: seed 2025). Significance brackets indicate the paired t-test p-value (vs. Local) and Cohen’s d effect sizes (vs. FedAvg, Freeze-FT).

Key observations:

1. **iPS-MFFL achieved the highest point estimate** for both final-round (85.42%) and best-round (85.52%) mean accuracy, though the one-way ANOVA across all methods did not reach statistical significance (p = 0.270), reflecting the small seed pool.
2. **Lowest variance.** iPS-MFFL’s SD (8.70 percentage points, pp) was the smallest among all methods; baseline SDs (11.77–21.09 pp) were 1.4× to 2.4× larger, suggesting **greater robustness** to partition heterogeneity (Figure 5b). Freeze-FT exhibited the highest variance (21.09 pp), suggesting its single-head approach is sensitive to the specific Dirichlet partition.
3. **Seed 42 resilience.** Under the most heterogeneous partition (seed 42; Table 2, Figure 5a), all methods suffered degraded performance. iPS-MFFL showed the smallest degradation: its seed 42 point estimate (75.38%) was 11.4 percentage points (pp) above the nearest FL baseline (FedAvg at 63.94%) and 25.8 pp above Freeze-FT (49.63%); aggregate performance across all three seeds remained 85.42 ± 8.70%.
4. **Consistent top performance.** On seeds 13 and 2025, iPS-MFFL achieved >90% accuracy, while all baselines remained below 88%.

**Table 1.** Dataset class distribution.

| Class | Training | Testing | Total |
| --- | --- | --- | --- |
| Glioma | 1,400 | 400 | 1,800 |
| Meningioma | 1,400 | 400 | 1,800 |
| Pituitary | 1,400 | 400 | 1,800 |
| No tumor | 1,400 | 400 | 1,800 |
| **Total** | **5,600** | **1,600** | **7,200** |

**Table 2.** Partition heterogeneity across seeds.

| Seed | Mean Client Entropy | Zero-class Instances | Characterization |
| --- | --- | --- | --- |
| 13 | 1.460 | 1 | Moderate heterogeneity |
| 2025 | 1.195 | 1 | High heterogeneity |
| 42 | 0.905 | 3 | Extreme heterogeneity |

### 3.2 Statistical analysis

#### Interpretation

The one-way ANOVA across methods did not reach statistical significance (p = 0.270), meaning we cannot formally reject the null hypothesis of equal means at α = 0.05. The pairwise comparisons in Table 5 are therefore reported as **exploratory analyses not protected by a significant omnibus test**, and should be interpreted cautiously. Nonetheless, the comparison with Local reached significance in a paired t-test (p = 0.013, d = 2.64), indicating a clear benefit of federated training over isolated training. Comparisons with FL baselines yielded medium-to-large effect sizes (d = 0.59–0.71) with achieved power of only 0.10–0.12 — far below the conventional 0.80 threshold. To detect effects of the observed magnitude with 80% power, **19–24 seeds would be required**. The non-significant p-values for FL baselines therefore reflect insufficient sample size (type II error risk) rather than evidence of equivalence, but they also cannot be interpreted as definitive evidence of superiority.

**Table 3.** Client-level class distribution for seed 42 (most heterogeneous partition).

| Client | Glioma | Meningio<br>ma | No tumor | Pituitary | Total | Entropy |
| --- | --- | --- | --- | --- | --- | --- |
| 0 | 68 | 0 | 766 | 0 | 834 | 0.41 |
| 1 | 1,298 | 1 | 462 | 82 | 1,843 | 1.06 |
| 2 | 26 | 718 | 0 | 136 | 880 | 0.81 |
| 3 | 4 | 250 | 171 | 918 | 1,343 | 1.23 |

| Client | Glioma | Meningioma | No tumor | Pituitary | Total | Entropy |
| --- | --- | --- | --- | --- | --- | --- |
| 4 | 4 | 431 | 1 | 264 | 700 | 1.02 |

**Table 4.** Test accuracy (%) across three random seeds. Final = accuracy at round 10; Best = maximum accuracy across all 10 rounds.

| Method | Seed 13 | Seed 42 | Seed 2025 | Mean Final<br>± Std | Mean Best |
| --- | --- | --- | --- | --- | --- |
| Local | 59.38 | 45.75 | 69.19 | 58.10 ±<br>11.77 | 58.10 |
| FedAvg | 87.00 | 63.94 | 84.50 | 78.48 ±<br>12.66 | 80.10 |
| FedProx | 87.31 | 61.44 | 86.25 | 78.33 ±<br>14.64 | 81.67 |
| Freeze-FT | 85.81 | 49.63 | 86.50 | 73.98 ±<br>21.09 | 79.75 |
| **iPS-MFFL** | **90.56** | **75.38** | **90.31** | **85.42 ± 8.70** | **85.52** |

**Table 5.** Pairwise comparison of iPS-MFFL vs. each baseline.

| Comparison | Mean $\Delta$ (pp) | t | p | Cohen's d | Power (post-hoc) | n for 80% power |
| --- | --- | --- | --- | --- | --- | --- |
| vs. Local | +27.31 | 8.74 | 0.013* | 2.64 | 0.66 | 4 |
| vs. FedAvg | +6.94 | 2.96 | 0.098 | 0.64 | 0.10 | 24 |
| vs. FedProx | +7.08 | 2.06 | 0.175 | 0.59 | 0.10 | 24 |
| vs. Freeze-FT | +11.44 | 1.60 | 0.251 | 0.71 | 0.12 | 19 |

\* $p < 0.05$ . One-way ANOVA across all five methods: $F = 1.52$ , $p = 0.270$ (not significant).

### 3.3 Convergence analysis

The round-by-round trajectory for seed 42 (**Figure 4**) illustrates the stability advantage of iPS-MFFL:

- **FedAvg, FedProx, and Freeze-FT** showed pronounced oscillation, with accuracy fluctuating by up to 25 pp between consecutive rounds (e.g., FedAvg: 67.1% at round 1 → 47.4% at round 3).
- **iPS-MFFL** exhibited smoother convergence with monotonic improvement after round 1, reaching 75.38% at round 10.

**Fig. 4.**
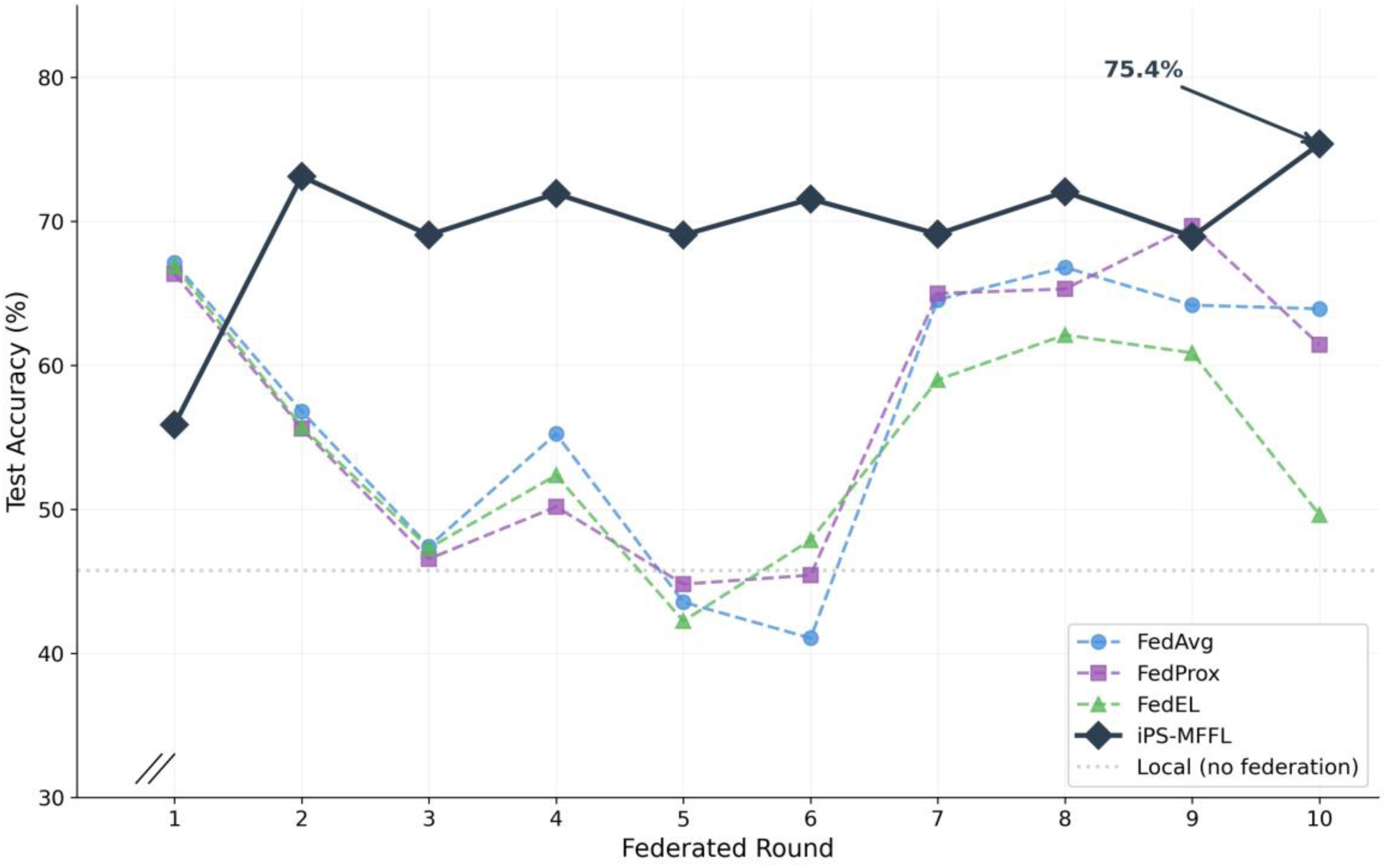
Round-by-round convergence under the most challenging partition (seed 42, H̅ = 0.91). iPS-MFFL (red) shows smoother convergence with monotonic improvement after round 1, while baselines exhibit pronounced oscillation due to conflicting local optima.

**Fig. 5.**
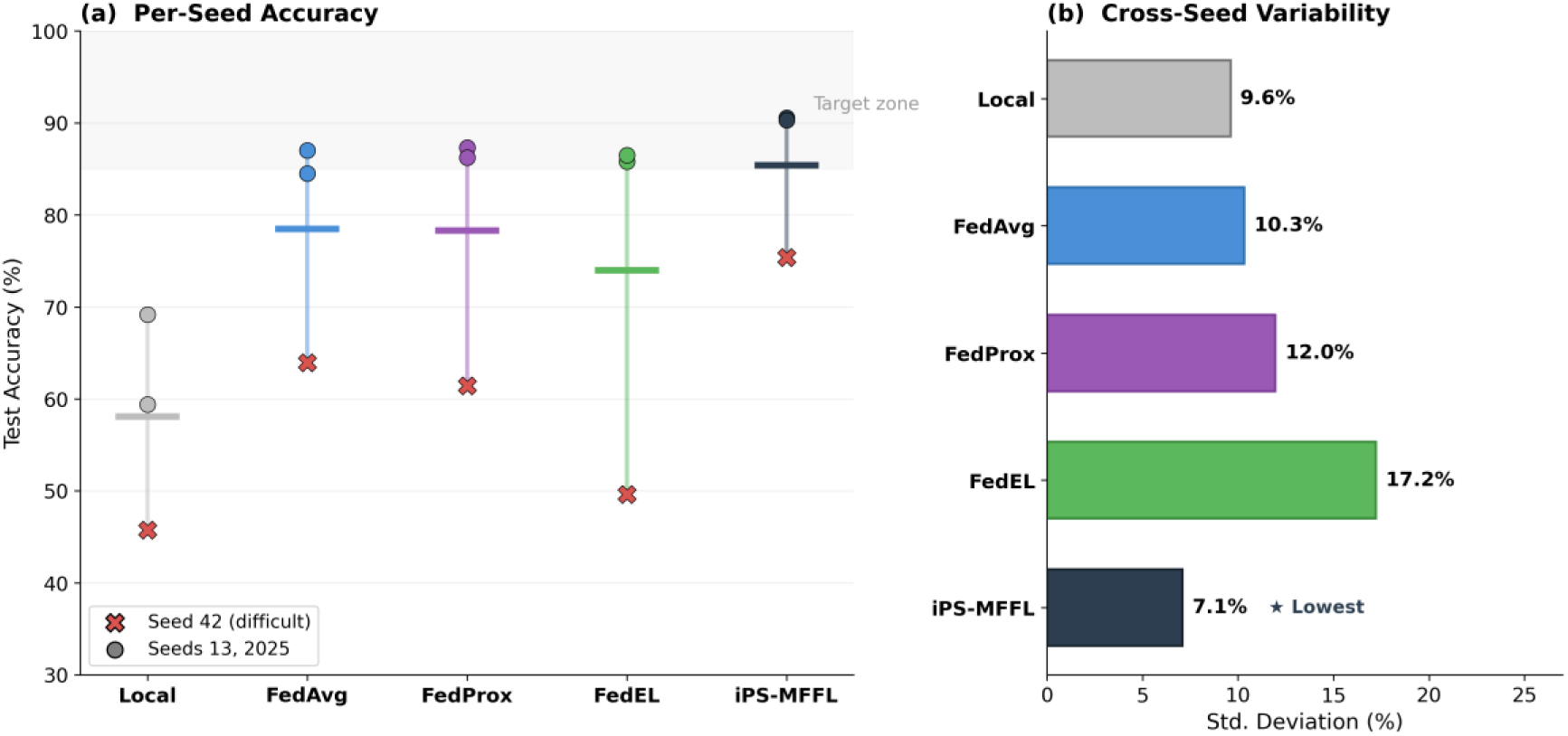
Robustness analysis. (a) Per-seed accuracy strip plot showing individual seed results (× = seed 42, extreme non-IID; ○ = seeds 13 and 2025) with mean lines. (b) Cross-seed standard deviation; iPS-MFFL achieves the lowest variance (8.7%), indicating the greatest robustness to partition heterogeneity.

This stability is consistent with the multi-head contrastive ensemble providing inherent regularization against the client drift that dominates under extreme non-IID conditions.

## 4. Discussion

### 4.1 Why federating weak learners independently helps under non-IID

The core insight of iPS-MFFL is that, once multiple weak-learner heads with heterogeneous training objectives are attached to a shared feature extractor, the way those heads are aggregated across clients becomes a distinct design choice. A single pooled aggregation collapses the diversity between heads and tends to average out the distinct decision boundaries that the heterogeneous objectives were designed to induce. Federating each weak learner at its own server preserves this diversity across rounds, and a separately federated meta-model learns to combine the diverse outputs in a way that reflects each client’s local data distribution.

Ensemble diversity theory [14] predicts that the ensemble error is bounded by the average individual error minus a term proportional to the diversity (disagreement) among learners. By aggregating each head independently, iPS-MFFL explicitly protects this diversity term at the federation layer — not only at the local training layer. The particular three-head configuration used in this study (see §2.7) is one concrete instantiation: each head is regularized in a complementary way (supervised signal, feature-space regularization via a model-contrastive term, and output-space regularization via an output-contrastive term), so that when one head is affected by client drift in a given round, the others can compensate.

We note that the independent contributions of the mcon and outcon heads have not been isolated in the current evaluation. While the Freeze-FT baseline (single sup head) serves as an ablation of the full multi-head system, determining whether outcon provides incremental value beyond mcon — or whether two heads would suffice — requires a factorial ablation that we leave to future work. The theoretical expectation is that feature-space (mcon) and output-space (outcon) regularization are complementary because they operate at different levels of the model hierarchy, but this remains to be empirically verified.

### 4.2 Robustness to partition heterogeneity

A central finding is that iPS-MFFL’s cross-seed SD (8.70 pp) was the smallest among all methods, with baseline SDs being 1.4× to 2.4× larger. The seed 42 partition was pathologically challenging (mean client entropy = 0.905, three zero-class instances) yet iPS-MFFL maintained 75.38% accuracy while Freeze-FT dropped to 49.63%. We note, however, that differences among methods did not reach statistical significance at the α = 0.05 level given the limited seed pool (n = 3).

We hypothesize that this robustness stems from iPS-MFFL’s hierarchical local-model architecture: independent aggregation of weak learners prevents a single catastrophic partition from corrupting all decision boundaries simultaneously, and the meta-model adaptively down-weights destabilized weak learners. This is supported by the observation that Freeze-FT — which shares the freeze-finetune principle but uses a single head — showed the highest variance (21.09%), suggesting that the single-head bottleneck amplifies partition sensitivity.

### 4.3 Benefit of ImageNet-pretrained ResNet18

Replacing a lightweight CNN with a pretrained ResNet18 provides: (1) richer 512-dimensional features with stronger visual semantics, (2) transfer learning that reduces the communication burden of federated rounds, and (3) more stable optimization via residual connections.

### 4.4 Computational overhead

The additional per-round computation of iPS-MFFL is modest: three fine-tuning passes instead of one, plus a lightweight meta-model step. Communication overhead is increased only for the three classification heads (3 × 512 × 4 = 6,144 weight parameters) and the meta-model (12 × 64 + 64 × 4 = 1,024 weight parameters), totalling 7,168 additional parameters per client — approximately 0.06% of the ResNet18 backbone (∼11.2M parameters) which remains shared.

### 4.5 The seed 42 phenomenon

Analysis of the seed 42 partition (Table 3) reveals why all methods struggled: Client 0 contains 91.8% “no tumor” images with zero meningioma and pituitary samples; Client 2 lacks “no tumor” entirely. This extreme heterogeneity causes all single-head methods to experience oscillatory convergence, as the global model alternately adapts to conflicting local optima. iPS-MFFL’s multi-head architecture mitigates this by maintaining distinct specializations that are combined adaptively.

### 4.6 Clinical implications

In clinical practice, the non-IID scenario tested here mirrors a realistic deployment: a consortium of hospitals ranging from large academic medical centers (with diverse case mixes) to small community clinics (handling primarily a few tumor types). A federated model must be robust to such heterogeneity. iPS-MFFL’s lower variance suggests it would provide more reliable performance across diverse clinical sites without site-specific tuning. A practical deployment scenario is a **triage system for rural or under-resourced hospitals** that see predominantly one tumor type but require a model capable of classifying all four categories when atypical cases present.

### 4.7 Limitations

1. **Small seed pool and non-significant omnibus test (n = 3).** While improving upon single-seed evaluation, n = 3 provides only 0.10–0.12 power for detecting the observed effect sizes (d ≈ 0.6) vs. FL baselines, and the one-way ANOVA did not reach significance (p = 0.270). Consequently, pairwise comparisons in Table 5 should be treated as exploratory rather than confirmatory. Future work should use ≥ 10 seeds (sufficient for 80% power at d = 1.0) or ideally ≥ 24 seeds (for d = 0.6); power calculations followed the standard non-central t-distribution formulation [15].
2. **No ablation study.** The current evaluation does not isolate the contributions of individual components (e.g., sup-only, sup+mcon, sup+outcon, all three; meta-model vs. majority voting). A full factorial ablation is planned to determine which components are load-bearing.
3. **Single dataset.** Validation on additional medical imaging datasets and the dataset’s community-curated provenance limit clinical generalizability.
4. **2D classification only.** Extension to 3D volumetric analysis and segmentation is a natural next step.
5. **Communication cost.** The three-server structure increases per-round communication for classifier parameters. A formal analysis is not provided.
6. **Hyperparameter sensitivity.** The current evaluation uses a single hyperparameter configuration (α = 0.3, K = 5, 10 rounds). Sensitivity analysis is needed.

## 5. Conclusions

We proposed iPS-MFFL, a federated learning framework in which (i) each client holds a shared feature extractor together with multiple weak-learner classification heads and a meta-model, and (ii) each weak learner and the meta-model are aggregated at **independent per-component servers** so that the diversity among weak learners is preserved across rounds. In our concrete instantiation we used an ImageNet-pretrained ResNet18 backbone, three weak learners trained with heterogeneous loss functions, and a small MLP meta-model. On a brain tumor MRI classification task under non-IID conditions (Dirichlet α = 0.3, K = 5 clients), iPS-MFFL achieved the highest point estimate of mean accuracy (85.42%) and the smallest cross-seed SD (8.70 pp) among all evaluated methods across three seeds. While differences among methods did not reach statistical significance in the one-way ANOVA (p = 0.270), the improvement over Local training was significant in a paired t-test (p = 0.013, d = 2.64), and medium-to-large effect sizes (d = 0.59–0.71) were observed relative to FL baselines. Post-hoc power analysis indicates that 19–24 seeds would be required for adequately powered inference. These results should be interpreted as preliminary evidence that combining transfer learning with structured weak-learner diversity and meta-learning is a promising approach to heterogeneous federated medical image classification, warranting further study. Future work should include expanded seed pools, ablation studies, additional baselines, multiple datasets, and real-world multi-institutional validation.

## Declarations

### Ethics statement

This study is a retrospective methodological investigation based on a publicly available, anonymized brain tumor MRI classification dataset. It does not involve recruitment of new patients, any intervention, or handling of identifiable personal data. The Brain Tumor MRI Dataset used in this study is publicly released on Kaggle [12] and does not require institutional ethics approval for secondary analysis. As a secondary analysis of fully anonymized, publicly available data, this study qualifies for exemption under the U.S. Common Rule (45 CFR 46.104(d)(4)) and is consistent with the principles of the Declaration of Helsinki (2013 revision), which permits research on anonymized data without individual informed consent when there is no risk to participants. No Institutional Review Board (IRB) review was sought because the data are already in the public domain and contain no identifiable information.

### Clinical trial registration statement

This study is **not an interventional study and does not constitute a clinical trial**. It is a retrospective methodological study using an existing, publicly available, anonymized dataset. Under the ICMJE definition, **clinical trial registration is not required**.

### Patient identifiability and de-identification statement

All imaging data originate from a **fully anonymized public dataset** (Brain Tumor MRI Dataset, Kaggle [12]). The data do not contain any of the HIPAA Safe Harbor 18 identifiers, specifically:

- No names, dates, telephone numbers, geographic data, Social Security numbers, medical record numbers, health plan beneficiary numbers, account numbers, certificate/license numbers, device identifiers, URLs, IP addresses, biometric identifiers, full-face photographs, or any other unique identifying number, characteristic, or code.

The authors confirm that this manuscript and its materials **do not create any risk of patient re-identification**.

### Patient and public involvement (PPI) statement

No direct Patient and Public Involvement took place at any stage of this research, including study design, conduct, reporting, or dissemination planning.

### Consent for publication

No new patient data or identifiable information is included; individual consent is not required.

### Distribution license

This preprint is distributed under a **Creative Commons Attribution 4.0 International (CC BY 4.0)** license (https://creativecommons.org/licenses/by/4.0/). Anyone may copy, distribute, transmit, and adapt the work for any purpose, including commercially, provided that appropriate credit is given, a link to the license is provided, and any changes are indicated.

### Data availability statement

The Brain Tumor MRI Classification dataset is publicly available on Kaggle (https://www.kaggle.com/datasets/masoudnickparvar/brain-tumor-mri-dataset). Source code will be released upon acceptance.

### Code availability statement

Python 3.11, PyTorch 2.3.1, CUDA 12.1.

## Acknowledgments

We thank Nickparvar for releasing the Brain Tumor MRI Classification dataset.

